# Advanced electrocardiography predicts computed tomography coronary artery calcium score

**DOI:** 10.1101/2025.03.05.25323202

**Authors:** Yash Bindal, Manoj Rajamohan, Daniel E Loewenstein, Todd Schlegel, Rebecca Kozor, Martin Ugander

## Abstract

**BACKGROUND:** Coronary artery calcification (CAC) has been linked to an increased risk of cardiovascular events. Its detection in asymptomatic individuals is valuable for reclassifying cardiac risk and informing management strategies. We hypothesised that an advanced electrocardiography (A-ECG) score derived from the standard 12-lead ECG predicts CAC with good diagnostic accuracy compared to computed tomography (CT).

**METHODS:** This retrospective study included patients that had undergone a 12-lead ECG and CT CAC scoring demonstrating either the absence (n=265) or presence (n=255) of any CAC. Multivariable elastic net logistic regression was used to generate an A-ECG score validated by nested resampling.

**RESULTS:** An A-ECG score for detecting CAC comprised of age, sex, and four ECG measures encompassing the duration of the Q wave in lead I, vectorcardiographic measures derived from the 12-lead ECG related to the spatial direction of the QRS complex (two measures) and the magnitude of the ST segment (one measure). Nested resampling estimated performance for predicting the presence of any CAC with an area under the receiver operating characteristics curve [95% confidence interval] of 0.78 [0.77-0.79], sensitivity 73 [72-75]%, specificity 66 [65-68]%, positive predictive value 70 [68-71]%, negative predictive value 71 [69-72]%, positive likelihood ratio 2.3 [2.1-2.4], and inverse negative likelihood ratio 2.6 [2.4-2.7].

**CONCLUSIONS:** The standard 12-lead ECG analysed by A-ECG analysis can predict the presence of CAC with good diagnostic performance. A-ECG may hold clinical utility as a low-cost and widely available initial screening test for the presence of CAC and cardiac risk prediction.

## Introduction

Coronary artery calcification (CAC) is a component of atherosclerosis and coronary artery disease (CAD). The presence of CAC is associated with adverse cardiovascular events^1^ and thus is used to help estimate cardiovascular risk in individuals and guide management. A measure of CAC through a calcium score is easy to acquire and interpret but involves a cost and a small amount of radiation. Efforts have been made predict the presence of CAC using alternative non-invasive non-radiographic investigations, such as electrocardiography (ECG).

Due to its mobility, low cost and widespread availability, ECG has experienced the introduction of advanced software-based techniques to improve its diagnostic value in measuring anatomical phenomena such as CAC through electrophysiological measures. However, current studies using ECG to detect CAC have had limitations. For example, ECG features have been combined with secondary clinical information such as systolic and diastolic blood pressures or known co-morbidities such as diabetes or overt CAD to develop models that sufficiently predict CAC^2^.

Alternatively, the probability of having any CAC has not directly been quantified from ECG measures, providing little clinical information about cardiovascular risk in comparison to calcium scoring^3^.

Advanced electrocardiography (A-ECG) techniques include three-dimensional vectorcardiographic measures derived from the standard 12-lead ECG via the Frank lead reconstruction technique of Kors, et al^4^, and waveform complexity measures derived from singular value decomposition (SVD)^5^. Compared to the conventional ECG, A-ECG has been shown to have higher diagnostic accuracy in conditions such as CAD, left ventricular hypertrophy, and left ventricular systolic dysfunction^6-9^. However, A-ECG has not yet been used to derive a score for predicting CAC. Therefore, the aim of this study was to create and test the utility of an A-ECG scoring system for predicting the presence of any CAC by CT.

## Methods

### Study design

This was a single-centre retrospective study between February 2017 and July 2023. It included low-to-intermediate risk chest pain patients assessed in an outpatient rapid access chest pain clinic who had a cardiac CT scan with CAC scoring and a 12-lead ECG. The study was approved by the local human research ethics committee (2019/STE10454 & 2019/ETH03639) and was approved with a retrospective waiver of consent.. Patients were excluded if they had ECG abnormalities that limited A-ECG analysis such as arrhythmias (e.g., atrial fibrillation), frequent ventricular ectopic beats, or QRS duration of >110 milliseconds.

### Data collection

Non-contrast cardiac CT scans were performed using ECG triggering on two clinical CT scanners (Philips iCT 256, Phillips Healthcare, Best, The Netherlands, and Canon Aquilion One Prism Edition, Canon Medical Systems, Tochigi, Japan, respectively). Cardiac CT scans were evaluated - no coronary calcification was defined as a CAC score of zero Agatston units (AU), and the presence of any calcification was defined as a CAC score of greater than zero AU. A clinical computerised 12-lead ECG system (Phillips Medical Systems, Best, The Netherlands) was used to acquire standard 10-second digital ECG files for A-ECG analysis. The patient inclusion process is illustrated in Figure 1.

**Figure 1:**
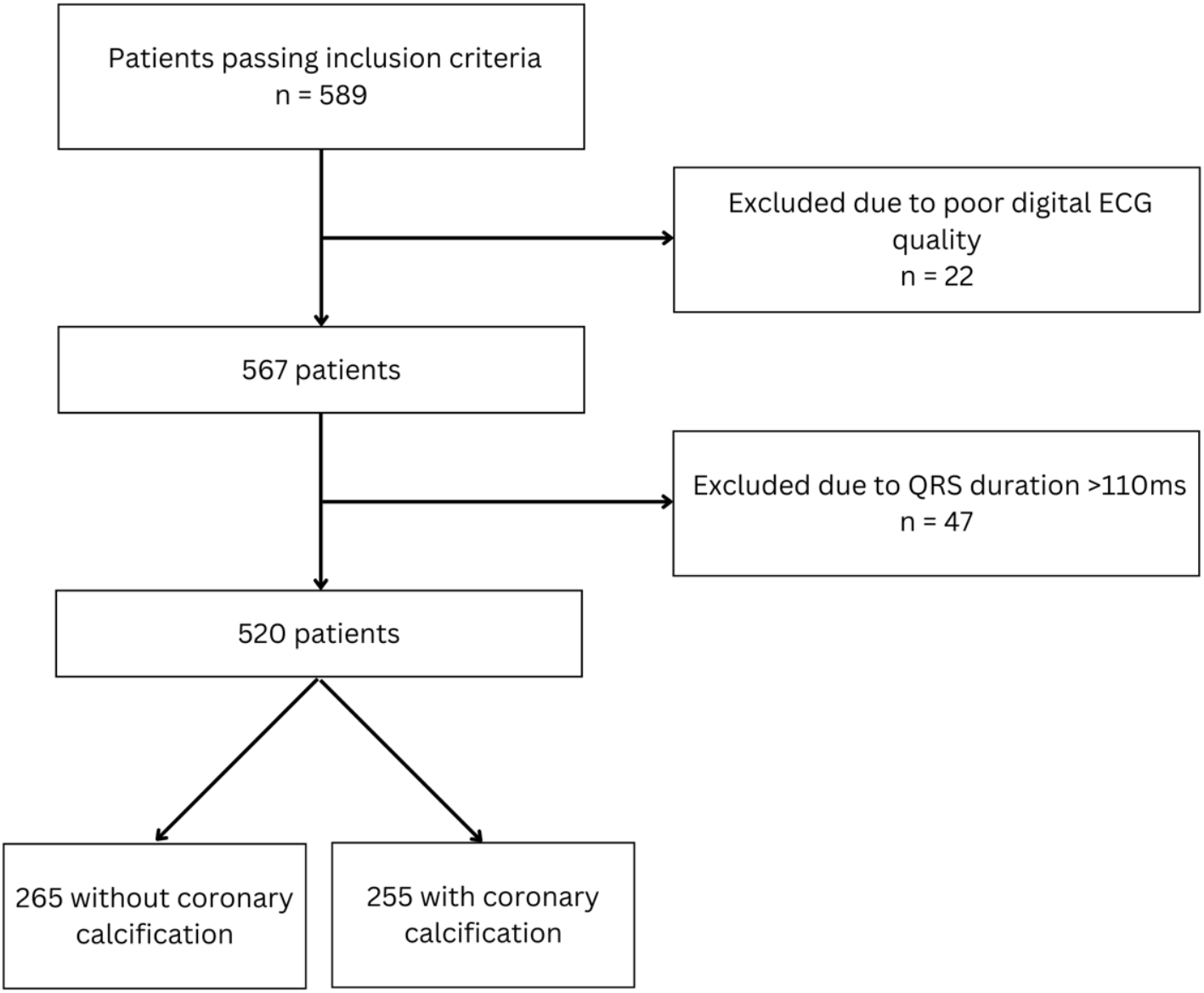
The patient inclusion flowchart. 589 patients met the inclusion criteria, after which 22 were removed from the study due to having poor ECG file quality which limited A-ECG analysis, and 47 were removed as they had a QRS duration greater than 110 milliseconds. Of the remaining 520 patients, 265 did not have coronary calcification while 255 did.

### Advanced ECG analysis

Analysis using A-ECG software was performed using in-house developed software^6,10^.Conventional ECG parameters were analysed in the software and included all major intervals, axes, and voltages. A-ECG parameters were obtained by signal averaging the 10-second ECG recording to generate values for parameters of: (1) derived three-dimensional ECG via the Frank lead reconstruction technique of Kors et al.^4^ to generate vectorcardiographic parameters, including the spatial mean and peaks QRS-T angle^11^, the spatial ventricular gradient, and various spatial waveform azimuths, elevations, and timevoltages^6^; and (2) QRS- and T-waveform complexity measures derived by singular value decomposition, for example the principal component analysis (PCA) ratio^10,12,13^ and the dipolar voltage equivalents of the QRS and T waveforms^6^. Most of the parameters studied have been described in other previous publications^6-9,14-16^.

### Statistical analysis

Statistical analysis was performed using freely available software (R version 4.3.1, R Core Team, R Foundation for Statistical Computing, Vienna, Austria) using packages dplyr^17^ for data transformation, tidymodels^18^ for nested resampling, and glmnet^19^ for logistic regression model building. Logistic regression was used to derive multivariable A-ECG scores for the probability of having any CAC, with a cut-off predicted probability of greater than 0.5. Elastic net regularisation was used with logistic regression to apply penalties to variable coefficients. This method was utilised as it enabled an optimal combination of Lasso and Ridge regularisation to produce a parsimonious A-ECG score. Previously, elastic net has outperformed Lasso and Ridge regularisation separately in terms of prediction accuracy^20^.

Hyperparameter tuning, model selection, as well as model evaluation was validated by using a nested resampling strategy. Using a nested resampling approach to estimate the generalization performance prevents information leakage between model derivation (hyperparameter tuning, model selection, and model fitting) and model evaluation. Since hyperparameter tuning and model selection is data-dependent, not including these steps in the resampling approach for model evaluation leads to optimistic, biased results^21,22^. This is avoided by using nested resampling which accounts for this added uncertainty since hyperparameters and model specification are not prespecified^21,22^. Nested resampling uses an outer loop of resampling for model evaluation and an inner resampling loop for hyperparameter tuning and model selection. Repeated stratified five-fold cross-validation was used in the outer loop, with each fold containing an approximately equal proportion of positive (calcium score greater than 0 AU) and negative cases (calcium score equal to 0 AU), the number of repeats were set to 10. In the inner loop, 100 bootstrap resamples were used to optimise the model hyperparameters, with tuning being performed by minimising the Brier Score^23^. For example, in the outer loop fold one was held out and folds two through five were used for the inner loop bootstrapping. These four folds were bootstrapped 100 times, and the model which averaged the best Brier score in 100 out-of-bag samples was selected to be fitted to fold one, the test set. This process was repeated four more times, which each one of folds two through five being used as the test set. Five sets of performance metrics (one from each fold) were combined and the entire process was repeated 10 times, yielding 50 sets of performance metrics. This process provided the expected performance of a best model configuration selected by 100 bootstraps in new, unseen data. Finally, 100 bootstraps were performed with the full dataset and the best model configuration was selected. The model, or A-ECG score, calculated the probability of having coronary calcification by logistic regression using the formula:

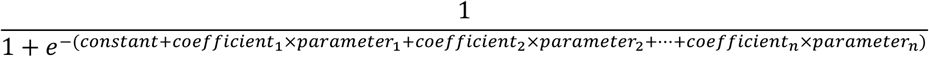

The variables and their coefficients of the best model configuration were compiled, and the patient values for each variable were collected.

## Results

### Study population

The study consisted of 520 patients, with 265 patients having no CAC (calcium score equal to 0 AU) and 255 patients having any CAC (calcium score greater than 0 AU). Table 1 shows the demographic characteristics of the cohort, split by presence of CAC. The group with no CAC was approximately one decade younger (no CAC 53±12 versus any CAC 65±12 years, p<001) and had fewer males (49.8% vs 60.0%) compared to the group with any CAC. The age of a male in the group without CAC was on average 12 years younger than a male with CAC (50±12 vs 63±11 years, p<001). A similar phenomenon was observed in female patients (no CAC 57±11 vs CAC 68±10 years, p<001).

**Table 1:**
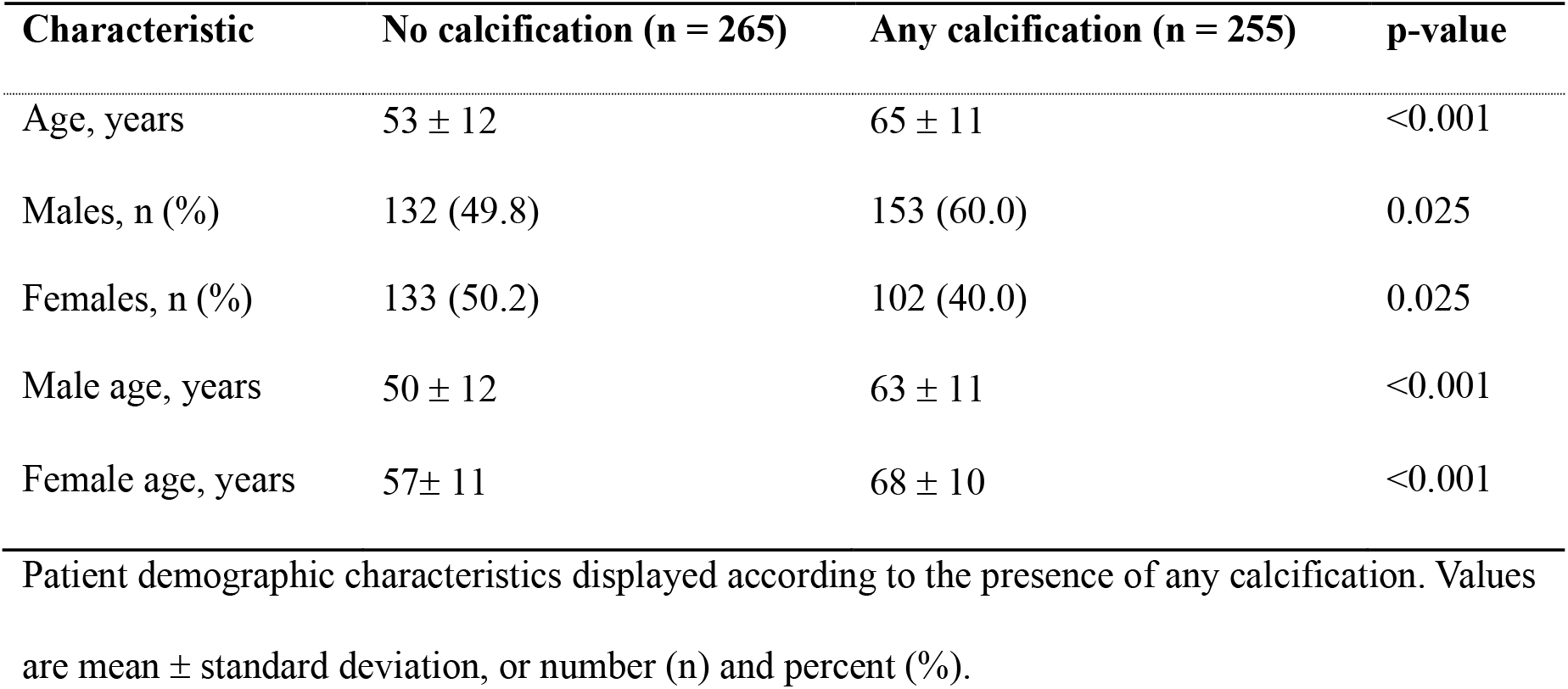
Demographic characteristics by coronary calcification status.

### Six-measure A-ECG score

The elastic net regularisation resulted in a penalty value of 0.04641589 and a mixture of entirely Lasso regularisation. The patient’s age at the time of calcium scoring and sex were identified as the most indicative factors for the distinction between no calcification and any calcification. The remainder of the score incorporated the following four ECG measures derived from the resting 12-lead ECG: (1) *Horizontal QRS 20*: the direction in degrees to which the three-dimensional QRS loop in the horizontal (or transverse) plane points at 20 milliseconds into that planar loop; (2) *QRS Elevation 3/8*: the elevation angle in degrees of the three-dimensional QRS loop when three eighths of the way into the loop; (3) *ST Magnitude 6/8*: the magnitude in microvolts of the three-dimensional ST segment loop when six eighths of the way into the loop; and (4) *Q Duration Lead I*: the duration of the Q wave in Lead I. The variables of the full six-measure A-ECG score and their coefficients can be found in Table 2 along with the patient values for each variable.

**Table 2:** Variables in the six-measure A-ECG score, split by coronary calcification status.

| Measure | Coefficient estimate | No calcification | Any calcification |
| --- | --- | --- | --- |
| Age, years | 0.06751267 | 53 ± 12 | 65 ± 11 |
| Male sex, n (%) | 0.401864 | 132 (50) | 153 (60) |
| Horizontal QRS 20, ° | -0.0000006581497 | 55 ± 36 | 46 ± 36 |
| QRS Elevation 3/8, ° | -0.004170926 | 30 ± 16 | 24 ± 15 |
| ST Magnitude 6/8, µV | -0.0002163557 | 327 ± 154 | 283 ± 142 |
| Q Duration Lead I, ms | -0.0116703 | 8 [0 – 12] | 6 [0 – 12] |
| Intercept | -3.991323 | - | - |

The variables and their coefficients of the final six-measure A-ECG score. The patients' values for each variable are shown, split by the presence of any calcification. Values are mean ± standard deviation for Age, Horizontal QRS 20, QRS Elevation 3/8, ST Magnitude 6/8. Values are total number (percent) for Sex. Values are the median [interquartile range] for Q Duration Lead I. °, degrees; µV, microvolts; ms, milliseconds.

Table 3 shows the performance of the score using nested resampling validation. Area under the receiver operating characteristics curve [95% confidence interval] was 0.78 [0.77-0.79], sensitivity 73 [72-75]%, specificity 66 [65-68]%, positive likelihood ratio 2.3 [2.1-2.4], and inverse negative likelihood ratio 2.6 [2.4-2.7].

**Table 3:** Performance measures of nested resampling models.

| Performance measure | Estimate | 95% Confidence Interval |
| --- | --- | --- |
| AUC | 0.78 | 0.77 – 0.79 |
| Sensitivity | 73% | 0.72 – 0.75 |
| Specificity | 66% | 0.64 – 0.68 |
| Accuracy | 70% | 0.69 – 0.71 |
| PLR | 2.27 | 2.13 – 2.42 |
| INLR | 2.56 | 2.42 – 2.70 |
| Brier score | 0.19 | 0.19 – 0.20 |

Median values and 95% confidence intervals of 50 sets of performance measures, obtained from the nested resampling procedure which had five hold-out folds and ten repeats. AUC, area under the receiver operating characteristics curve; INLR, inverse negative likelihood ratio; PLR, positive likelihood ratio

## Discussion

This study shows that an A-ECG score including age, sex and four A-ECG variables, can predict the presence of any CAC with good diagnostic performance.

### Age, sex, and CAC

Patient age and sex were the two non-ECG measures included in the final A-ECG score (Table 2). Previous literature has considered clinical information such as blood pressure and known diagnosis of CAD when attempting to predict presence of CAC, measures which are not always available to the physician^2^. Alternatively, age and sex are measures which are readily available when performing an ECG, and do not require additional medical examinations or history.

Age and sex had the highest magnitude of contribution onto the A-ECG score, which is in agreement with the current knowledge surrounding the pathogenesis of CAC. This is also seen in a previous study specifically evaluating the power of clinical and ECG measures in predicting CAC^2^. Patients with calcification were older on average, and this is consistent with existing literature which shows that the prevalence of any detectable calcification increases as age increases in both males and females^24,25^. Additionally, the difference in the mean ages of females and males was 8 and 6 years in the no CAC and any CAC groups, respectively. This is an observation similar to existing literature which suggests that CAC prevalence is seen at an earlier age in males than in females, with the median level of CAC occurring 10 years earlier in life^25^. The associations of both age and sex with CAC have been widely observed before^24-26^, validating the use of these two measures in the final A-ECG score to achieve optimal diagnostic accuracy.

### Advanced ECG measures

Four ECG features were included in the final score. The vectorcardiogram derived from the resting 12-lead ECG describes the instantaneous spatial orientation and magnitude of electrical activity over the cardiac cycle, something which is not immediately apparent using the conventional ECG.

The A-ECG measure *QRS Elevation 3/8* refers to the elevation angle in degrees of the vectorcardiographic three-dimensional QRS loop when three eighths of the way into the loop. Elevation is the angle between the VCG vector and the horizontal plane, with angles directed inferiorly having a positive value. In patients with no calcification, the mean *QRS Elevation 3/8* value was 30 degrees. Similar values are seen in a previous study using ‘normal’ male subjects admitted to hospital for reasons other than cardiovascular disease^27^. In the group with detectable calcification, the mean value was 24 degrees, illustrating that patients from this group had, on average, a more horizontal angle that the QRS loop made with the horizontal plane three eights of the way through the loop. A similar phenomenon has been reported previously, with less elevation of the QRS loop in post myocardial infarct (MI) subjects with ventricular tachycardia compared to healthy controls^28^. In principle, a QRS loop closer to the horizontal plane in the diseased group could be suggestive of hypertrophy of the anterior portion of the left ventricle, or ischemia of the inferior myocardium, causing an upwards-directed shift of cardiac electrical activity. Although CAC may not have specific clinical manifestations, it is associated with an increased risk of coronary events and is highly prevalent in patients with CAD^1^. The mechanism of how both CAC and CAD cause ischemia and hypertrophy is widely accepted, and the coexistence of CAC and cardiac hypertrophy can cause a greater coronary event risk^29^.

The A-ECG measure of *Horizontal QRS 20* refers to the direction in degrees to which the QRS loop in the horizontal plane points at 20 milliseconds into that loop. In patients with no CAC, the mean value was +55 degrees, whereas it was +46 degrees in the group with any CAC. This more leftward and/or posterior initial QRS vector in our patients with CAC has also been noted in left and right ventricular hypertrophy, ventricular tachycardia after MI, or anterior wall myocardial fibrosis caused by coronary artery disease, all disease states which can be linked to CAC^28,30-33^.

The A-ECG measure *ST Magnitude 6/8* refers to the magnitude in microvolts of the three-dimensional ST segment loop when six eighths of the way into the loop. The ST segment loop extends from the end of the QRS wave to the end of the T wave and is represented by the vector which connects these two endpoints. In patients without and with CAC, the mean ST Magnitude 6/8 values were 327 and 283 microvolts, respectively, with earlier studies also having noted associations between increasing age and decreasing mean ST segment loop magnitude in ‘normal’ male patients^27,34^. These values represent the magnitude six eighths of the way through the ST segment loop, which is approximately localised to the zone between the T wave upstroke and T peak given that three eighths, four eighths, and five eighths interval measurements of the ST segment loop have been shown to represent the transition between the ST segment and the T wave upstroke^34^. A lower mean magnitude of the ST segment loop in the zone between the T wave upstroke and T peak, as observed in the group with CAC, could be suggestive of lower amplitudes, or flattening, of T waves which has been shown to be associated with myocardial ischemia or electrolyte abnormalities^35^. Sufficient narrowing of the coronary vessels due to CAC can induce ischemic changes in the myocardium, which in turn is detectable by ECG and VCG changes^36^.

The A-ECG measure of *Q Duration Lead I* refers to the duration of the Q wave in Lead I, from the first negative deflection following the P wave through to the beginning of the R wave. The Q wave in Lead I represents the normal left-to-right depolarisation of the interventricular septum and thus, small septal Q waves can be expected in lead I. In patients with no CAC, the median value for this measure was 8 milliseconds. In patients with any CAC, the median value was shorter at 6 milliseconds. A shorter Q wave duration in lead I could be suggestive of a thinner interventricular septum, or less initial rightward forces due to LVH which has previously been linked to CAC. However, these differences are of a small magnitude, especially considering that the median values for both groups in the cohort fall outside the generally accepted criteria for pathological Q waves of greater than or equal to 30 milliseconds.

Taken together, the four A-ECG measures in the final model have a pathophysiological association, either via previous studies or through known pathophysiological principles with structural changes in cardiovascular diseases in which CAC is a significant component.

### Performance

Through logistic regression with elastic net regularisation and nested resampling, performance of multiple best models was collected. With an accuracy of 70%, sensitivity of 73% and specificity of 66% (Table 3), a correct classification of no CAC or any CAC could be expected to be achieved in just under three-quarters of cases. The reported likelihood ratios allow an interpretation of the expected performance of the A-ECG score at an individual level, in a manner that is independent of the prevalence of disease. For example, the median inverse negative likelihood ratio suggests that following a negative result as determined by an optimal A-ECG score, a patient is 2.6 times less likely to have CAC as determined by CT. Overall, the performance through this approach shows that A-ECG has potential in serving as an inexpensive and rapid tool for identifying CAC. However, just over one-quarter of subjects could have a false negative or false positive result.

### Practical application

Looking forward, A-ECG may hold promise for various clinical applications where low-risk, inexpensive, and rapid screening tools can serve to provide initial risk stratification. This is particularly significant in informing the decision to progress to more definitive but costly and higher-risk investigations such as calcium scoring or CT coronary angiography, both of which are not always readily available in all clinical situations. On the contrary, ECG is inexpensive and broadly available, and often routinely conducted in patients being investigated for cardiac disease. Furthermore, a derived diagnostic A-ECG score, like the much familiar calcium score, provides a numeric probability value of the presence of any CAC which is available rapidly after conducting the ECG. This would facilitate immediate discussion between physicians and patients regarding the need for lifestyle modifications, preventative therapy, and follow-up investigations to reduce the risk of CAC and, in general, promote cardiovascular health.

### Limitations and future directions

This was a single-centre study, with the cohort comprised of patients referred for outpatient cardiac assessment. Thus, it is likely that both groups in this population contain a higher prevalence of cardiac co-morbidities than what would be expected in the general population. The applicability of the derived A-ECG score would benefit from validation in cohorts from different centres.

Patients with atrial fibrillation and conduction delay were excluded from the study and therefore, the A-ECG score developed could not be applied to patients with these conditions without further study in such populations. This is particularly noteworthy considering that CAC is commonly prevalent in patients with atrial fibrillation^37,38^.

The modelling strategy used in the current study was internally validated through nested resampling. However, before being applied in a clinical decision scenario the derived A-ECG score needs to be externally validated with evaluation of calibration-in-the-large, discrimination slope, as well as its clinical usefulness with decision-curve analysis, and such studies are justified.

Finally, attempts to predict the presence or degree of purely any anatomical phenomenon via electrophysiology are limited. Differences in estimations of disease via imaging versus electrophysiology may indicate different manifestations of underlying anatomical versus electrical disease processes^39^. Moreover, it is not entirely clear that anatomical measures such as CAC scoring necessarily serve as optimal “gold standards” for measures of electrophysiology, the latter providing different and complementary information and having its own independent predictive utility^2,40^.

## Conclusion

A six-measure score derived from A-ECG analysis of the conventional 12-lead ECG had good diagnostic performance for predicting the presence of CAC by CT. A-ECG score holds potential clinical utility and value as a low-cost and widely available tool, but requires further large clinical trials to establish outcomes.

## Data Availability

All data produced in the present work are contained in the manuscript

